# Association of Hippocampus volume with normal serum Natrium levels and predictive analyses of cognitive adversities in non-demented middle-aged and older adults

**DOI:** 10.1101/2024.11.01.24316554

**Authors:** Asma Hallab, Alzheimer’s Disease Neuroimaging Initiative

## Abstract

**Introduction:** Serum Natrium abnormalities are largely observed in older adults and are associated with higher risks. Less is known about the association between serum Natrium variations and medial temporal brain structures, mainly involved in cognition and memory. The study’s objective was to explore the association between serum Natrium and Hippocampus volume and to assess associated cognitive risks.

**Methods:** Non-demented ADNI3 participants (healthy controls (HC) and with mild cognitive impairment (MCI)) with complete serum Natrium, ADAS_13_ score, and Hippocampus volume at baseline were included. Linear and non-linear associations were evaluated. To assess the odds of MCI, logistic regression adjusted was performed. Holm method was used to adjust for Family-wise error rate in the main analysis and reported as a *q*-value.

**Results:** A total of 469 cases with a median age of 70 years (IQR: 66, 76) were included. The median serum Natrium level was 141 (IQR: 139, 142). Serum Natrium levels showed a significant association with Hippocampus volume in the total study population and MCI subgroup (Adj.*ß*_MCI_=-95 (−162, −28), *p*=0.006, *q*=0.036). Serum Natrium levels did not show a significant association neither with the ADAS13 total score (Adj.*ß*_Total_=-0.04(−0.28, 0.21), *p*=0.8) nor with the odds of being diagnosed with MCI at baseline (OR_MCI_= 1.00(0.88, 1.13), *p*= 0.935).

**Conclusions:** Normal Serum Natrium variations were significantly associated with Hippocampus volumes depending on the underlying neurodegenerative pathology, thus, without predicting clinically relevant cognitive adversity. Further studies are needed to better understand the mechanisms and assess protective factors.

**Key findings:**

- Serum Natrium levels within normal ranges were significantly associated with Hippocampus volume.
- The association between Serum Natrium levels and Hippocampus volume was particularly significant in participants with mild cognitive impairment.
- There was no significant association between serum Natrium and ADAS_13_ total score.
- Serum Natrium levels within normal ranges did not predict concomitant risk of mild cognitive impairment.

## 1. Introduction

Natrium is a key electrolyte that plays a pivotal role in human homeostasis through maintaining volume balance, neural signaling, and muscular contractions. (1) A clinically relevant dysregulation of its levels is associated with severe manifestations and leads to life-threatening risks. (1) Older adults represent a particular risk group with a higher prevalence of pathological Natrium levels, particularly Hyponatremia. (2) Etiologies might vary between endogenic factors (such as the syndrome of inappropriate antidiuretic hormone secretion (SIADH), the renal salt wasting syndrome (RSW), (3) or kidney disease) (4) and exogenous factors (mainly iatrogenic or nutritional). (1)

Hyponatremia is associated with higher morbidity and mortality risks. (5) Affected old patients are more exposed to frailty, falls, fractures, and depressive mood, (6, 7) and those with higher frailty scores have a higher prevalence of electrolyte abnormalities, particularly Hyponatremia. (8) In patients with underlying severe pathologies, Hyponatremia is associated with worse outcomes. (9) Furthermore, hyponatremic states might be associated with elevated epileptic cerebral activities in electrophysiological studies and with clinical seizures. (10) While Hyponatremia is largely studied, older patients are also exposed to Hypernatremia with associated risks. (11)

Studies on the relationships between Hyponatremia and cognitive dysfunction showed controversial results. Some studies reported a significant association with cognitive decline in older adults, either impacting overall cognition or executive function, (12) while others found no significant associations. (13) Delirium, an acute form of cognitive dysfunction commonly reported in older persons, is also significantly associated with Hyponatremia. (14, 15) While the effect of serum Natrium on brain functions is supported by rodent (16, 17) and clinical studies, (12) a very limited number of studies explored the relationship between cognition-relevant brain structures of the medial temporal lobe and serum Natrium levels.

The main aim of this work was to study the association between serum Natrium levels within normal ranges and the Hippocampus volume, a main biomarker of cognitive function, in non-demented ≥ 50 years adults. The secondary aim was to evaluate whether a non-pathological variation in serum Natrium levels might modulate the overall cognition scores or the odds of cognitive impairment in advanced ages.

## 2. Methods

### 2.1. Study population

ADNI 3 study participants without dementia were included in the study. Alzheimer’s Disease Neuroimaging Initiative (ADNI) is a longitudinal observational cohort performed in different centers across the USA and Canada. Dr. M. Weiner is the principal investigator, and the project is NIH-funded (U.S. National Institute on Aging: Grant U19 AG024904 - more in the declaration of funding and acknowledgment). ADNI3 is the fourth phase (following ADNI1, ADNIgo, and ADNI2), during which all study participants underwent 3 Tesla magnetic resonance imaging (MRI). The study was conducted in accordance with the declaration of Helsinki. Ethical approval was obtained from each local recruitment center’s IRB. Participants gave written consent. The current study is a secondary analysis of anonymized data and respects ADNI’s data use agreement (DUA). Further information, study protocols, and ethical approvals can be found at https://adni.loni.usc.edu.

### 2.2. Serum Natrium

Serum Natrium levels were assessed at baseline in the ADNI3 cohort. Values were reported in mmol/L. Missing values and defect measurements were checked manually and removed. In case of duplicate measurement, both values were checked, and only the first one was kept as long as a therapeutic intervention between both measurements cannot be verified. Normal clinical values were equal to or between 135 and 145 mmol/L.

### 2.3. Magnetic resonance imaging

All MRIs performed during ADNI3 phases were 3 Tesla. Segmentation was performed using FreeSurfer 6.0 (https://surfer.nmr.mgh.harvard.edu). The total Hippocampus volume was studied (UCSF version – cross-sectional analysis), and was calculated as the sum of the volume of the right and left corresponding structures for each volume of interest (VOI). The intracranial volume was also adjusted for in the regression model as a strong predictor of Hippocampus variation. Detailed protocols of ADNI3 neuroimaging methods in different ADNI recruitment sites can be found at https://adni.loni.usc.edu/adni-3/.

### 2.4. Overall cognition

Overall cognition was estimated based on the total Alzheimer’s Disease Assessment Scale – Cognitive Subscale – 13 items (ADAS_13_) score at baseline. For a larger description of the cognitive status of the study population, information on the total Clinical Dementia Rating - Sum of Boxes (CDR-SB) was included.

### 2.5. Inclusion criteria

ADNI 3 study participants with dementia (n=83), without serum Natrium measurements (n= 10), or with serum Natrium levels beyond pathological clinical cutoffs (n=18), without VOI (n= 102), without complete ADAS_13_ total score at baseline (n= 5), and missing information on age (n= 3) were excluded. Overall, 469 healthy controls (HC) and mild cognitive impairment (MCI) cases were included.

### 2.6. Statistical analysis

Continuous variables were reported as median (interquartile ranges: IQR) and count / categorical variables as number (%). Differences between groups were assessed using the Wilcoxon rank sum test, Pearson’s Chi-squared test, and Fisher’s exact test, depending on the variable’s type, and the corresponding *p*-value was reported each time. ADAS_13_ explores overall cognition, while Hippocampus volume was considered a relevant neuroimaging biomarker for cognition. First, the correlation between age, serum Natrium levels, neuropsychological test scores, and VOI was visualized in a matrix to assess the relationship between different variables and exclude eventual multicollinearity. Then, the association between Hippocampus volume as a dependent variable and serum Natrium level (mmol/L) as an independent continuous variable were explored in linear regression models. Afterward, relevant confounding factors, such as age, sex, educational level (years), main diagnosis (HC and MCI), ADAS_13_ total score, APOE ε4 status (none, one, or two alleles), and the period between blood sampling and MRI (days) were adjusted for. Non-linearity was explored using restricted cubic splines based on different knots (3, 4, and then 5). The same strategy was followed after cognition-related stratification and models were adapted to HC- and MCI-strata (removing cognition-related diagnosis as a confounder in the model).

The secondary analysis was based on the linear regression with the ADAS_13_ total score as the dependent variable, following the same strategy used for Hippocampus volume.

Then, logistic regression was used to assess the odds of MCI diagnosis based on serum Natrium variation. First, a crude model was explored; then, it was adjusted for the same relevant confounding variables (age, sex, educational level (years), ADAS_13_ total score, APOE ε4 status, and the period between blood sampling and cognitive testing (days)). Non-linearity was tested using the same method, and corresponding models were visualized. The odds ratio (OR), 95% confidence interval (*CI*), and corresponding two-sided *p*-value were reported. To avoid type I errors related to stratification and modeling, the Holm-Bonferroni method was used to adjust for the family-wise error rate (FWER) and reported as a two-sided *q*-value. The significance level for both *p* and *q-*values was set at 0.05. RStudio version 2024-04.1-748 was used for the statistical analysis and data visualization.

## 3. Results

### 3.1. Description of the study population

The median age was 70 years (IQR: 66, 76), and HC were significantly younger than MCI participants (69 vs. 73, *p*-value < 0.001), 257 were females (55 %), and 212 were males (45 %). The median ADAS_13_ score at baseline was 10 points (IQR: 6, 14), and the median Hippocampus volume was 7,510 mm^3^ (IQR: 6,946, 8,106) (Table 1).

**Table 1:** Characteristics of the study population and comparison between diagnostic groups.

| *Characteristics* | *N* | *Total study population*<br>*N = 469<sup>1</sup>* | *Healthy controls*<br>*N = 298<sup>1</sup>* | *Mild cognitive impairment*<br>*N = 171<sup>1</sup>* | *p-value<sup>2</sup>* |
| --- | --- | --- | --- | --- | --- |
| **Age (years)** | 469 | 70 (66, 76) | 69 (66, 74) | 73 (67, 77) | **<0.001** |
| **Sex** | 469 |  |  |  | **<0.001** |
| Female |  | 257 (55%) | 186 (62%) | 71 (42%) |  |
| Male |  | 212 (45%) | 112 (38%) | 100 (58%) |  |
| **Racial profile** | 469 |  |  |  | 0.12 |
| White |  | 381 (81%) | 234 (79%) | 147 (86%) |  |
| Black |  | 58 (12%) | 41 (14%) | 17 (9.9%) |  |
| Other |  | 30 (6.4%) | 23 (7.7%) | 7 (4.1%) |  |
| **Educational level (years)** | 469 | 16.00 (16.00, 18.00) | 17.00 (16.00, 18.00) | 16.00 (15.50, 18.00) | 0.3 |
| **Marital status** | 469 |  |  |  | 0.12 |
| Currently married |  | 354 (75%) | 218 (73%) | 136 (80%) |  |
| Currently not married or unknown |  | 115 (25%) | 80 (27%) | 35 (20%) |  |
| **Housing situation** | 469 |  |  |  | >0.9 |
| House or apartment |  | 451 (96%) | 287 (96%) | 164 (96%) |  |
| Retirement or nursing institution |  | 14 (3.0%) | 8 (2.7%) | 6 (3.5%) |  |
| Other |  | 4 (0.9%) | 3 (1.0%) | 1 (0.6%) |  |
| **APOE ε4 status (numbers)** | 365 |  |  |  | **0.016** |
| 0 allele |  | 230 (63%) | 161 (67%) | 69 (56%) |  |
| 1 allele |  | 113 (31%) | 71 (29%) | 42 (34%) |  |
| 2 alleles |  | 22 (6.0%) | 9 (3.7%) | 13 (10%) |  |
| Missing values |  | 104 | 57 | 47 |  |
| **Hippocampus volume (mm<sup>3</sup>)** | 469 | 7,510 (6,946, 8,106) | 7,616 (7,068, 8,180) | 7,228 (6,583, 7,747) | **<0.001** |
| **ADAS<sub>13</sub> score (points)** | 469 | 10 (6, 14) | 8 (5, 11) | 14 (11, 19) | **<0.001** |
| **CDR-SB (points)** | 469 | 0.00 (0.00, 1.00) | 0.00 (0.00, 0.00) | 1.00 (0.50, 2.00) | **<0.001** |
| **Serum Natrium level (mmol/L)** | 469 | 141.00 (139.00, 142.00) | 141.00 (139.00, 142.00) | 141.00 (139.00, 142.00) | >0.9 |
| **Period between Blood sample and MRI (days)** | 469 | 10 (6, 20) | 11 (6, 18) | 10 (6, 22) | 0.8 |
| **Period between Blood sample and ADAS<sub>13</sub> (days)** | 469 | 36 (25, 62) | 33 (23, 61) | 41 (26, 63) | 0.2 |

<sup>1</sup> Median (IQR); n (%), <sup>2</sup> Wilcoxon rank sum test; Pearson's Chi-squared test; Fisher's exact test
**ADAS<sub>13</sub>:** Alzheimer's Disease Assessment Scale – Cognitive Subscale 13 items, **APOE:** Apolipoprotein E, **CDR-SB:** Clinical Dementia Rating – Sum of Boxes, **MRI:** Magnetic Resonance Imaging.

The median serum Natrium level was 141 (IQR: 139, 142). Serum Natrium showed no statistically significant difference between different groups, with a median equal to 141 mmol/L in HC and MCI groups (Figure 1a).

**Figure 1:**
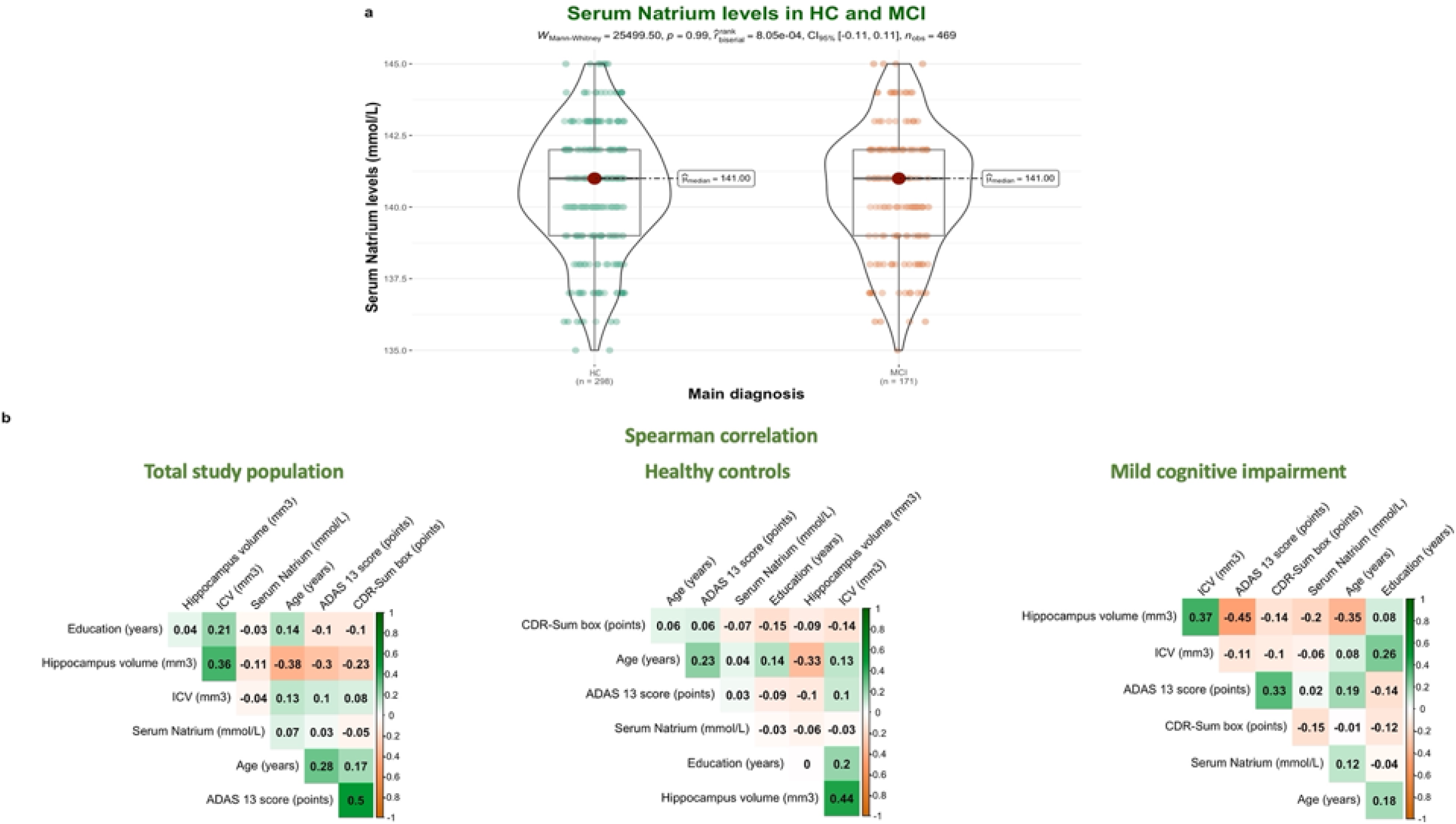
Visualization of data exploration**. Figure 1a:** Difference in serum Natrium levels between healthy controls and mild cognitive impairment**. Figure 1b:** Spearman correlation matrix in the total study population and different strata**. ICV:** Intracranial volume, **HC:** Healthy controls, **MCI:** Mild cognitive impairment.

### 3.2. Correlation analysis

The Spearman correlation did not show any high correlations between the different predictors used for the analysis (Figure 1b).

### 3.3. Association between serum Natrium and Hippocampus volume

In the total study population, the association between Hippocampus volume and serum Natrium showed significant results in the linear model (Crude *ß*= −53 (−92, −14), *p*-value _linear_ = 0.008, *q*-value _linear_ =0.036 & Adj. *ß*= −43 (−77, −8.9), *p*-value _linear_ = 0.013, *q*-value _linear_ =0.036). The variations associated with an increase of one mmol/L in serum Natrium are equivalent to −0.71% (crude model) and −0.57% (adjusted model) of the median Hippocampus volume in the total study population. After stratifying for the cognition-related diagnosis, no significant associations were found in HC, while in the MCI group, an increase of one mml/L in serum Natrium level was associated with a decrease of 103 mm^3^ in Hippocampus volume (*ß*= −103 (−175, −30), *p*-value _linear_ = 0.006, *q-*value _linear_ =0.036). After adjusting for relevant confounding factors, an increase in Natrium levels of one mmol/L was significantly associated with a decrease of 95 mm^3^ in Hippocampus volume (Adj. *ß _MCI_*= −95 (−162, −28), adj. *p*-value _linear_ = 0.006). These variations are equivalent to −1,43% (crude model) and −1,31% (adjusted model) of the median Hippocampus volume in MCI. After adjusting for FWER, the observed *p*-values remained statistically significant (*q*-value = 0.036). Non-linear models did not show a statistically significant superiority over the linear model.

Linear models are summarized in Table 2 and visualized in Figure 2.

**Table 2:**
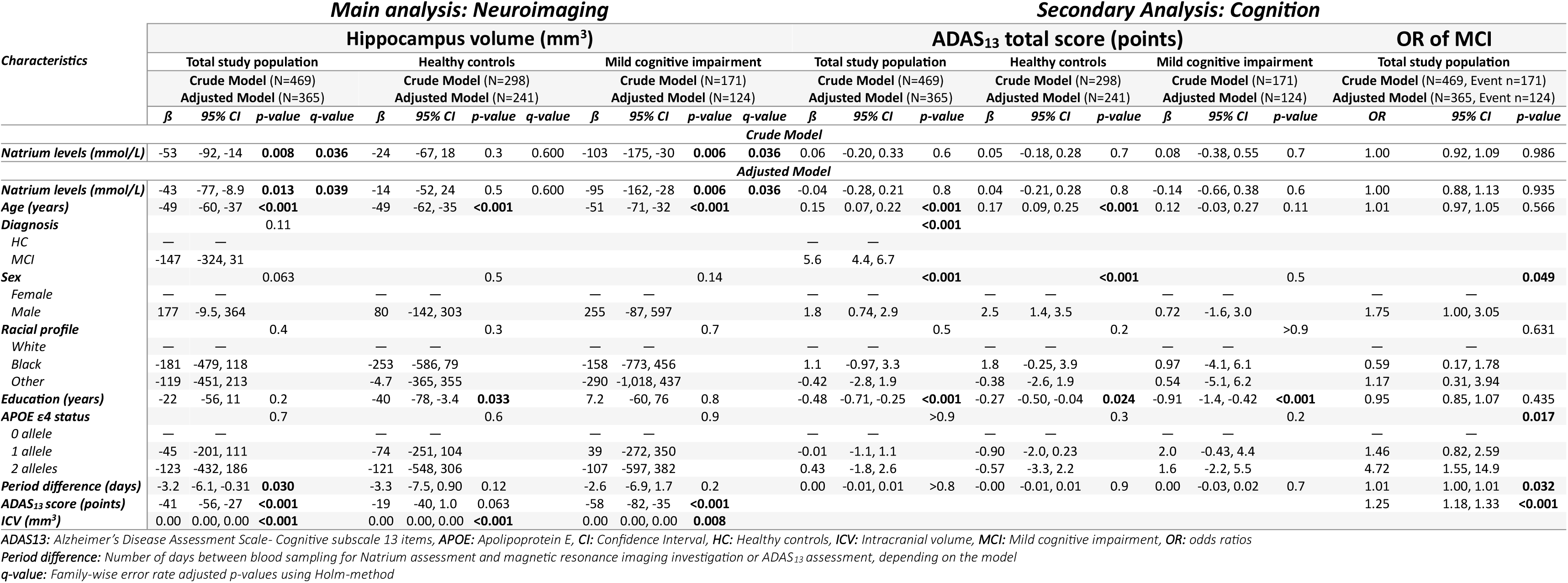
A summary table of linear and logistic regression models related to serum Natrium associations with neuroimaging and cognition.

**Figure 2:**
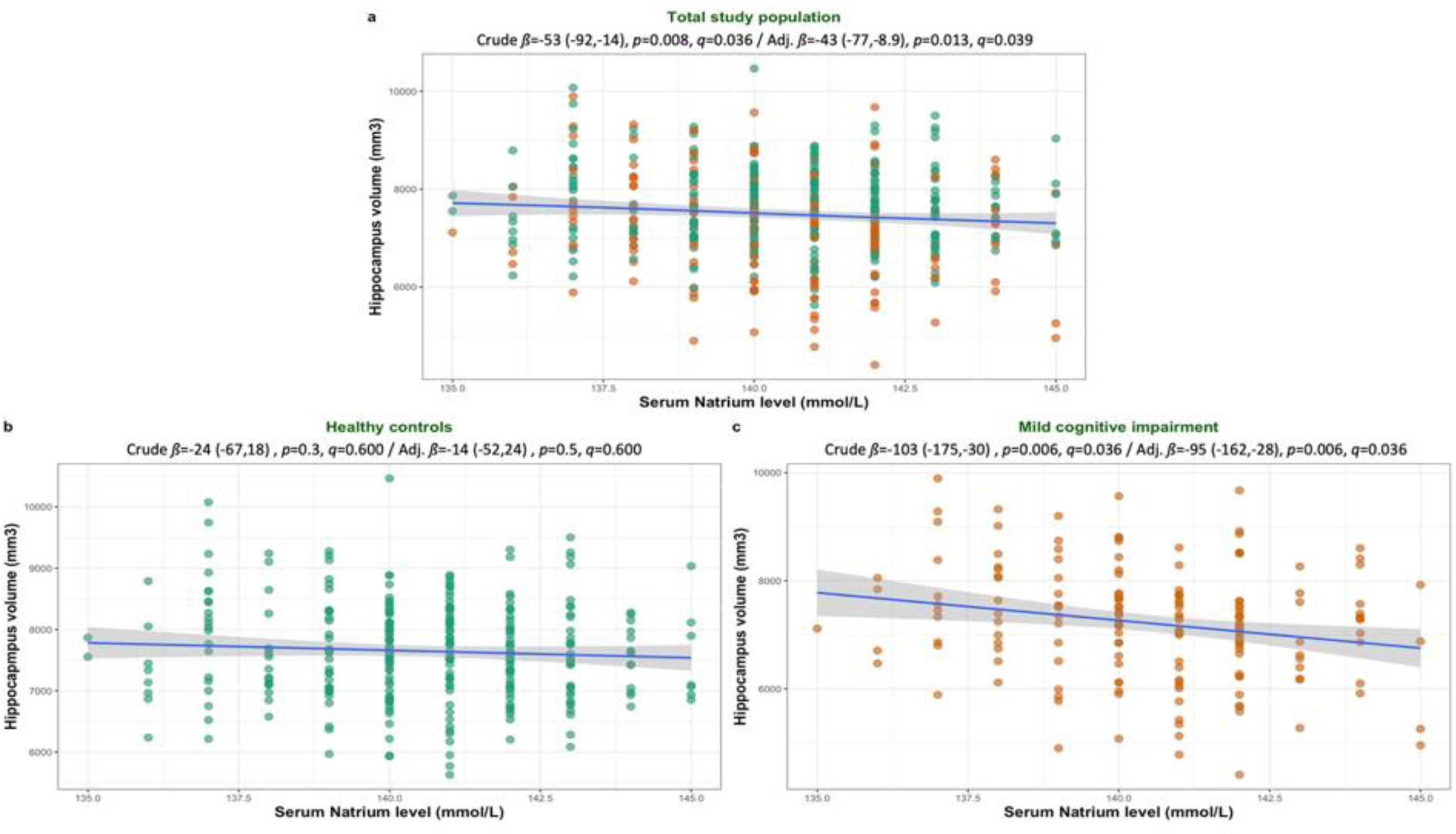
Association between serum Natrium levels and Hippocampus volumes**. Figure 2a:** Total study population**. Figure 2b:** Healthy controls**. Figure 2c:** Mild cognitive impairment**. *q*-value:** Family-wise error rate adjusted *p-*value using the Holm-Bonferroni method.

### 3.4. Association between serum Natrium and cognitive decline

#### ADAS_13_ total score

No statistically significant linear association between ADAS_13_ scores and serum Natrium levels was found in the total study population (*ß*= 0.06 (−0.20, 0.33), *p*-value _linear_ = 0.6). Neither adding a non-linear factor to the model nor stratifying for the cognition-related diagnosis did improve the model prediction. Similarly, adjusting for confounding factors did not show significant associations (*ß*= −0.04 (−0.28, 0.21), *p*-value _linear_ = 0.8). The linear models are detailed in Table 2 and visualized in Figure 3a.

**Figure 3:**
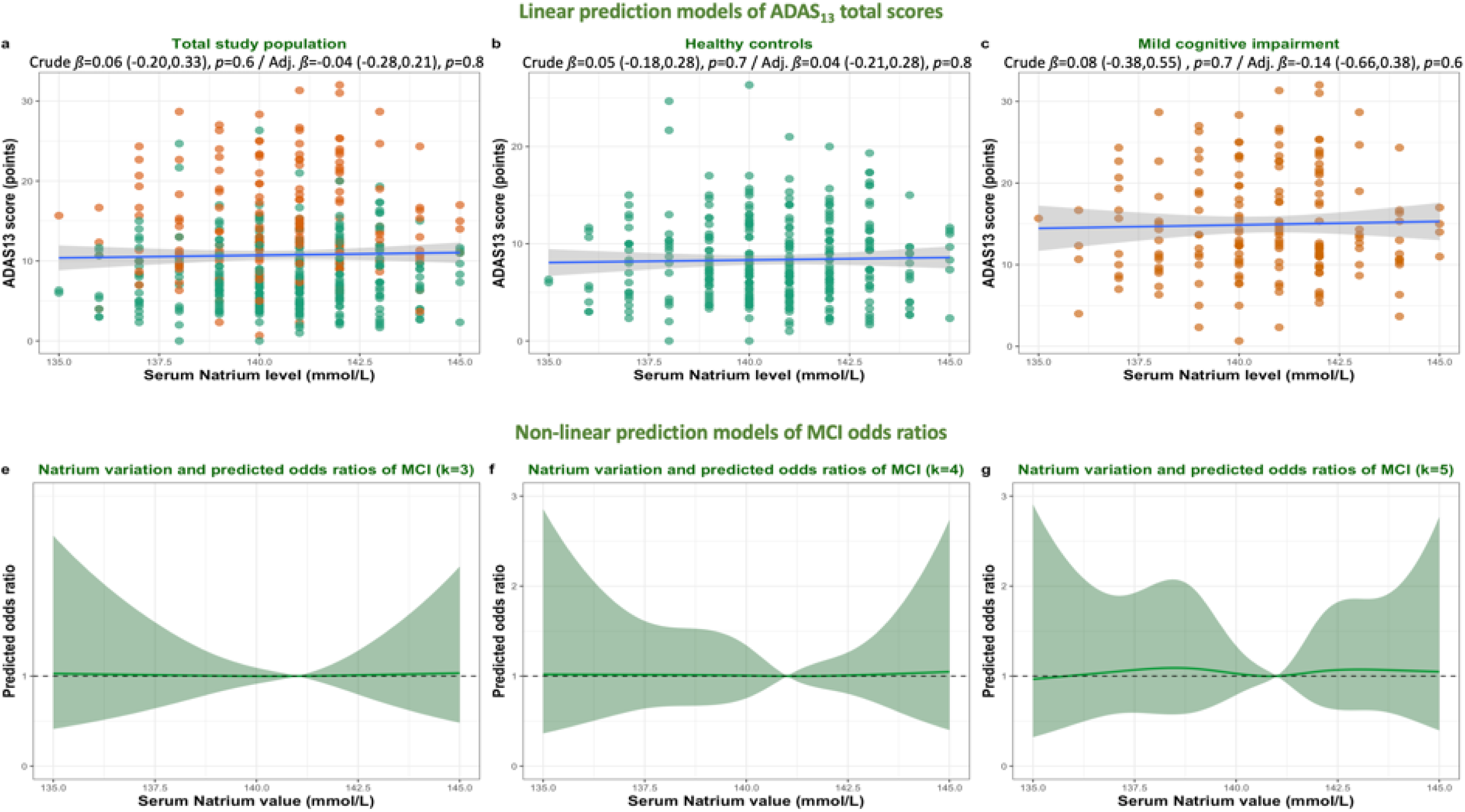
Association between serum Natrium levels and cognitive adversities**. Figure 3a:** Linear association with ADAS13 total score in the total study population and different strata**. Figure 3b:** Non-linear models on the predicted odds ratios of mild cognitive impairment using different knots**. ADAS_13_:** Alzheimer’s Disease Neuroimaging Initiative – Cognitive subscale - 13 items, **MCI:** Mild cognitive impairment.

#### Mild cognitive impairment

In logistic regression, no significant association was found between serum Natrium levels and the odds of being diagnosed with MCI at baseline, neither in a crude linear model (OR= 1.00 (0.92, 1.09), *p-*value _linear_ = 0.986) nor after adjustment for confounders (adj. OR= 1.00 (0.88, 1.13), adj. *p*-value _linear_ =0.935) (Table 2). Non-linear models did not show any significant predicted odds ratios (Figure 3b).

## 4. Discussion

This study presented additional evidence on the association between serum Natrium levels in non-demented adults and Hippocampus volume. The main outcome was the significant linear association between Hippocampus volume and normal serum Natrium levels in MCI, but not HC.

A recent longitudinal study on patients with Hyponatremia found a significant correlation between medial-temporal structures and Natrium volume changes. (18) A correction of Hyponatremia was significantly correlated with decreased Hippocampus volume, which might be explained by water retention during Hyponatremia in vulnerable structures such as the Hippocampus. Those results were based on a longitudinal analysis but limited by a low number of patients with Hyponatremia.

Neurodegeneration observed in MCI patients might present a risk factor for higher sensitivity to serum Natrium variations. Rodent studies showed that during the very early stages of Alzheimer’s disease, the hippocampal astrocyte-endothelium physical interaction is lost, and the vascular density is significantly reduced compared to patients without AD pathology. This mechanism is followed by disruption of the blood-brain barrier and leakage of tracers in rodent models. (19) Furthermore, high Natrium intake leads to systemic tissue Natrium excess, and this form of subclinical edema is associated with aging. (20)

Although no significant associations between ADAS_13_ and serum Natrium levels were found in the current analysis, previous studies reported significant associations between hyponatremic states and results of MMSE, DemTect, CERAD, and Trail-Making Test A and B (TMT-A and TMT-B). (12, 18) A study on community-dwelling older men showed a significant association between cognitive impairment (assessed with modified-Mini-Mental-Status (3MS) and TMT-B) and serum Natrium levels in the first and third tertials (126-140 mmol/L and 143-153 mmol/L, respectively) over a median follow-up period of 4.6 years. (21) Another study found that the association between serum Natrium levels and cognitive scores was non-linear, and only serum Natrium levels below 135 mmol/L were significantly associated with cognitive decline. (22)

The absence of statistically significant associations might be related to excluding study participants with serum Natrium levels outside the clinical normal ranges. Those might have affected cognition at a clinically measurable level. Moreover, the cross-sectional analysis does not exclude longitudinal associations in the case of chronic non-corrected Hyponatremia. Natrium measurements are only provided at baseline in ADNI3, and a very low number of ADNI participants had Natrium levels slightly beyond normal cutoff values and did not meet the inclusion criteria.

No association between baseline serum Natrium levels and the odds of being diagnosed with MCI at baseline was found. In a cross-sectional analysis of people receiving peritoneal dialysis, Hyponatremia was significantly associated with cognitive impairment, particularly executive dysfunction. (9) In contrast, in a longitudinal analysis of a larger population-based scale, the hazard ratio of dementia incidence over 10.7 years was not associated with Natrium levels. (13) Cerebrovascular diseases were reported as risk factors for incident dementia in patients with Hyponatremia, and the severity of Hyponatremia might increase the risks, as patients with severe Hyponatremia had a hazard ratio (HR) of 4.29 to develop dementia, while the HR in those with non-severe Hyponatremia was 2.08. (23)

The delay between blood sampling and MRI was not a significant predictor in the MCI-specific adjusted model. Although serum Natrium levels might show constant variations between short measurement intervals, its variation beyond normal limits is generally related to serious pathological states. This might explain the absence of influence of the delay between the two explorations on the statistical significance of the model.

A precautious screening of Hyponatremia in older patients, particularly those admitted to healthcare structures, is recommended to ensure prompt interventions and prevent harmful consequences. A correction of the hyponatremia was associated with an improvement in neurocognitive tests in randomized controlled trials. (24, 25) The reversibility of the neuropathological disruptions and clinical symptoms related to Hyponatremia, particularly on Hippocampus structures and correlated cognitive manifestations, (16) should motivate careful monitoring and correction of Natrium levels in older adults and vulnerable groups.

### Strengths

The association between serum Natrium and Hippocampus volume was rarely studied despite the clinical evidence in favor of an association between serum Natrium variations and cognition. This study explored a situation where serum Natrium levels were within normal range. The results highlight an eventual vulnerability of the Hippocampus microstructure during aging, particularly at the MCI stage. Although the analysis found no significant association between serum Natrium levels and cognition, the population might reflect a “pre-symptomatic” stage of cognition-specific adversities associated with Hippocampus vulnerability to serum Natrium variability. Previous studies have shown that Natrium molecules impact local immune reactions and tissue homeostasis. (26) The strength of this study lies in its ability to pave the way for further research, particularly by raising new questions on the brain-blood barrier (BBB) and Natrium’s association with neuroinflammation biomarkers in the Hippocampus tissue at MCI stages. Pathophysiological investigations within at-risk groups and over a longitudinal timeframe seem crucial.

### Limitations

The first limitation of this study is the low number of included participants compared to the whole ADNI cohort. This is mainly due to the availability of serum Natrium levels only in ADNI3. Larger studies are needed to better explore subgroups.

The second limitation is related to the very low number of participants with clinically pathological serum Natrium levels in ADNI3, which had to be excluded from the current analysis since there is no way to control for serum osmolality in the case of pathological Natrium values and therapeutic intervention between different explorations can not be verified. It is thus noteworthy that ADNI3 has strict inclusion criteria, and participants with higher morbidity risks were not eligible. Therefore, it is assumed that included subjects are less likely to present a severe underlying condition interfering with serum Natrium levels or severely impacting its variability. Nevertheless, it seems crucial to explore neuroimaging data in larger populations with chronic Natrium dysregulations to extend the risk assessment.

The third limitation is related to missing information on medication that might modulate serum Natrium levels. This missing information does not seem to impact the current study question, where the main exposure was serum Natrium levels, independently from underlying etiologies. By focusing on one simple, measurable, and clear exposure, the introduction of recall- and information bias, which are generally common in large-scale cohorts, was avoided.

## 5. Conclusions

Serum Natrium plays an important role in maintaining stable cognitive function in middle-aged and older adults, and its variation presents a significant association with changes in volumetric biomarkers of cognition, particularly the Hippocampus volume. The association is modulated by several factors, mainly the underlying cognitive and neurodegenerative state at the MCI stage. Having a serum Natrium level within clinical normal ranges seems – on its own – not sufficiently protective against Hippocampus volume variations. Thus, the changes do neither seem to affect the overall cognition nor the concomitant risk of MCI. Larger studies are needed to better understand the underlying mechanisms and their eventual associations with impaired BBB integrity and to address adequate therapeutic management in case of pathological Natrium levels to slow or revert eventual associated damage.

## Declarations

### Data availability

All data used in the manuscript is available at https://adni.loni.usc.edu.

### Ethical approval and consent

ADNI was performed in accordance with the Declaration of Helsinki. Ethical approval was obtained from local IRBs corresponding to each ADNI’s recruitment center. All participants gave written consent. The current study is based on a secondary analysis of anonymized data and respecting ADNI’s data use agreement (DUA).

### Author contributions

AH has full access to all of the data and takes responsibility for the integrity of the data and the accuracy of the analysis, visualization, drafting, and editing of the manuscript.

## Acknowledgments

“Data collection and sharing for the Alzheimer’s Disease Neuroimaging Initiative (ADNI) is funded by the National Institute on Aging (National Institutes of Health Grant U19 AG024904). The grantee organization is the Northern California Institute for Research and Education. In the past, ADNI has also received funding from the National Institute of Biomedical Imaging and Bioengineering, the Canadian Institutes of Health Research, and private sector contributions through the Foundation for the National Institutes of Health (FNIH) including generous contributions from the following: AbbVie, Alzheimer’s Association; Alzheimer’s Drug Discovery Foundation; Araclon Biotech; BioClinica, Inc.; Biogen; Bristol-Myers Squibb Company; CereSpir, Inc.; Cogstate; Eisai Inc.; Elan Pharmaceuticals, Inc.; Eli Lilly and Company; EuroImmun; F. Hoffmann-La Roche Ltd and its affiliated company Genentech, Inc.; Fujirebio; GE Healthcare; IXICO Ltd.; Janssen Alzheimer Immunotherapy Research & Development, LLC.; Johnson & Johnson Pharmaceutical Research &Development LLC.; Lumosity; Lundbeck; Merck & Co., Inc.; Meso Scale Diagnostics, LLC.; NeuroRx Research; Neurotrack Technologies; Novartis Pharmaceuticals Corporation; Pfizer Inc.; Piramal Imaging; Servier; Takeda Pharmaceutical Company; and Transition Therapeutics.”

